# Sex disparity in acute myeloid leukemia – evidence from a study of *FLT3*-ITD mutated patients

**DOI:** 10.1101/2020.09.04.20188219

**Authors:** Caroline Engen, Monica Hellesøy, Tim Grob, Bob Löwenberg, Peter J.M. Valk, Bjørn T. Gjertsen

## Abstract

Little attention has been directed at untangling sex-related molecular and phenotypic differences in AML. While increased incidence and poor risk is generally associated with a male phenotype, *FLT3*-ITD, *NPM1* and *DNMT3A* mutations are overrepresented in female AML. Here, we have investigated the relationship between sex and *FLT3*-ITD mutation status by comparing clinical data, mutational profiles, gene expression and *ex vivo* drug sensitivity responses in four cohorts: the Beat AML cohort, the LAML-TCGA cohort and two independent HOVON/SAKK clinical trial-associated cohorts, comprising a total of 1755 AML patients. We found that sex-associated molecular differences were prevalent in *FLT3*-ITD mutated AML. Co-occurrence of *FLT3*-ITD, *NPM1* and *DNMT3A* mutations was overrepresented in females, while males with *FLT3*-ITDs were characterised by additional mutations in genes involved in RNA splicing and epigenetic modification. Female and male *FLT3*-ITD mutated AML had diverging expression of multiple leukemia-associated genes, as well as discrepant *ex vivo* drug-responses, suggestive of discrete functional properties. Surprisingly, we found significant prognostication of *FLT3*-ITD only in female patients. Thus, we suggest optimisation of *FLT3-*ITD mutation status as a clinical tool in a sex-adjusted manner. We further hypothesize that prognostication, prediction and development of therapeutic strategies in AML can be improved by including sex-specific considerations.

## INTRODUCTION

Sex influences regulatory mechanisms of the haematopoietic system as well as innate and adaptive immune responses.^1,2^ Strong age- and sex-specific discrepancies in incidence of autoimmune conditions^3^ and cancer,^4^ including hematological malignancies,^5^ suggest fundamental genetic and endocrine sex-related variability.

Androgens have been used to treat various bone marrow (BM) failure syndromes since the 1960s,^6^ and the presence of hormone receptors on hematopoietic cells, including malignant cell populations, has been recognised for decades.^7^ Yet, little is known about the molecular mechanisms modulating hematopoiesis through sex hormone pathways, or their contribution in hematopoietic malignancies. It has been indicated that sex-hormone receptors significantly contribute in regulation of hematopoietic cell subsets, including stem and progenitor cells.^8^ Sex-disparity in acute myeloid leukemia (AML) incidence is known, with a progressive male excess with increasing age.^9^ It has also been shown that male AML patients have significantly inferior outcomes compared to females, both in adult and in pediatric AML.^10^ Sex-specific mutational profiles in AML have also been described; *FLT3*-ITD, *NPM1* and *DNMT3A* mutations are overrepresented in females,^11,12^ while mutations in *RUNX1, ASXL1, SRSF2, STAG2, BCOR, U2AF1* and *EZH2* are more prevalent in males.^12^ Female overrepresentation among AML patients with co-occurrence of *DNMT3A, NPM1* and *FLT3*-ITD mutations has also been reported.^13^

Despite the demographic differences in survival and somatic mutation profiles, sex-specific considerations are currently not made in therapeutic assessment or clinical risk stratification. Amongst the mutations with reported sex-dependent discrepancies is internal tandem duplications (ITD) of the FMS-like tyrosine kinase 3 (*FLT3*). *FLT3*-ITD is present in approximately 25% of AML cases, and is a negative prognostic marker that is integrated in standard risk stratification guidelines in AML as well as guiding FLT3-targeted therapy.^11,14^ Yet, the association between sex and *FLT3*-ITD mutations has not been explored in depth. Here, we present results from genomic, functional and time-to-event-analyses of four well-annotated cohorts, including the Beat AML cohort,^15^ the LAML-TCGA cohort^16^ and two independent HOVON/SAKK cohorts, in sum comprising 1755 AML patients. Cohort compositions with regards to sex and *FLT3* mutation status is described in Table 1, and comparative analyses of sex-differences related to clinical parameters are presented in Supplementary tables 1-3.

**Table 1.** – Composition of all cohorts in relation to sex, age and FLT3 status.

|  |  |  | Beat AML | HOVON1 | HOVON2 | LAML-TCGA |
| --- | --- | --- | --- | --- | --- | --- |
| **Total** | All | Number | 496* | 432 | 625 | 200 |
|  |  | Median age (range) | 61 (2-87) | 46 (15-77) | 53 (18-65) | 57 (18-88) |
|  | Female | Number | 222 | 215 | 273 | 92 |
|  |  | Median age (range) | 57.5 (2-85) | 46 (16-77) | 53 (19-65) | 57 (21-88) |
|  | Male | Number | 276 | 217 | 352 | 108 |
|  |  | Median age (range) | 63 (5-87) | 46 (15-75) | 55 (18-65) | 58 (18-83) |
| **ITD+** | All | Number | 123 | 117 | 146 | 39 |
|  |  | Median age (range) | 61 (10-85) | 47 (18-77) | 50 (18-65) | 57 (22-83) |
|  | Female | Number | 62 | 67 | 74 | 18 |
|  |  | Median age (range) | 60 (22-79) | 45 (18-77) | 53 (19-65) | 58.5 (25-68) |
|  | Male | Number | 61 | 50 | 72 | 21 |
|  |  | Median age (range) | 61 (10-85) | 47.5 (19-71) | 48.5 (18-65) | 57 (22-83) |
| **ITD-** | All | Number | 373 | 315 | 479 | 161 |
|  |  | Median age (range) | 62 (2-87) | 45 (15-75) | 55 (18-65) | 57 (18-88) |
|  | Female | Number | 160 | 148 | 199 | 74 |
|  |  | Median age (range) | 55.5 (2-85) | 46 (16-74) | 52 (19-65) | 57 (21-88) |
|  | Male | Number | 213 | 167 | 280 | 87 |
|  |  | Median age (range) | 64 (5-87) | 45 (15-75) | 65 (18-65) | 58 (18-81) |

\*The sample selection analysed from the Beat AML cohort comprises a total of 498 samples, but age for two of these samples is not annotated

## METHODS

### Patient sample selection

We analysed four independent patient cohorts; The Beat AML, LAML-TCGA, HOVON 1 and HOVON 2 cohorts. Beat AML and LAML-TCGA cohorts were used for descriptive analyses of somatic variant profiles. The Beat AML cohort was further investigated by differential gene expression (DGE) analyses, and exploration of *ex vivo* drug response profiles. All four cohorts were included for time-to-event analyses. Data from Beat-AML^15^ and LAML-TCGA^16^ are publicly available. All patients in the two HOVON cohorts provided written informed consent at trial inclusion.

The Beat AML cohort comprises 672 specimens from 562 individuals. We restricted our analysis to samples the All.variants.csv file, downloaded from http://www.vizome.org/aml/geneset/ on the 01.11.2018. This included 608 samples from 519 individuals. At inclusion, 15 individuals had two samples acquired from different tissues. The sample with the lowest number of recurrent mutations was discarded. We filtered the remaining samples based on the variable “dxAtSpecimenAcquisition” and retained samples annotated as “Acute myeloid leukaemia (AML) and related precursor neoplasms”, resulting in a total of 571 samples. For descriptive analysis of somatic variants, we included only the first sample when serial samples were present, resulting in 498 samples from 498 individuals. For DGE and drug response, we analysed 390 samples from 360 individuals and 359 samples from 322 individuals, respectively, including only samples with sample IDs overlapping with the exome sequencing data set (Supplementary figure 1A). For outcome assessment, we restricted the analysis to diagnostic samples, denoted “Initial Acute Leukemia Diagnosis”, where survival data was present, resulting in 303 individuals (Supplementary figure 1B). Notably, we constructed an extended *FLT3*-ITD annotation based on the combination of the consensus *FLT3*-ITD variable from the clinical summary table (Table S5-Clinical Summary) and Pindel call of *FLT3*-ITDs, as reported in the All.variants.csv file.

**Figure 1.**
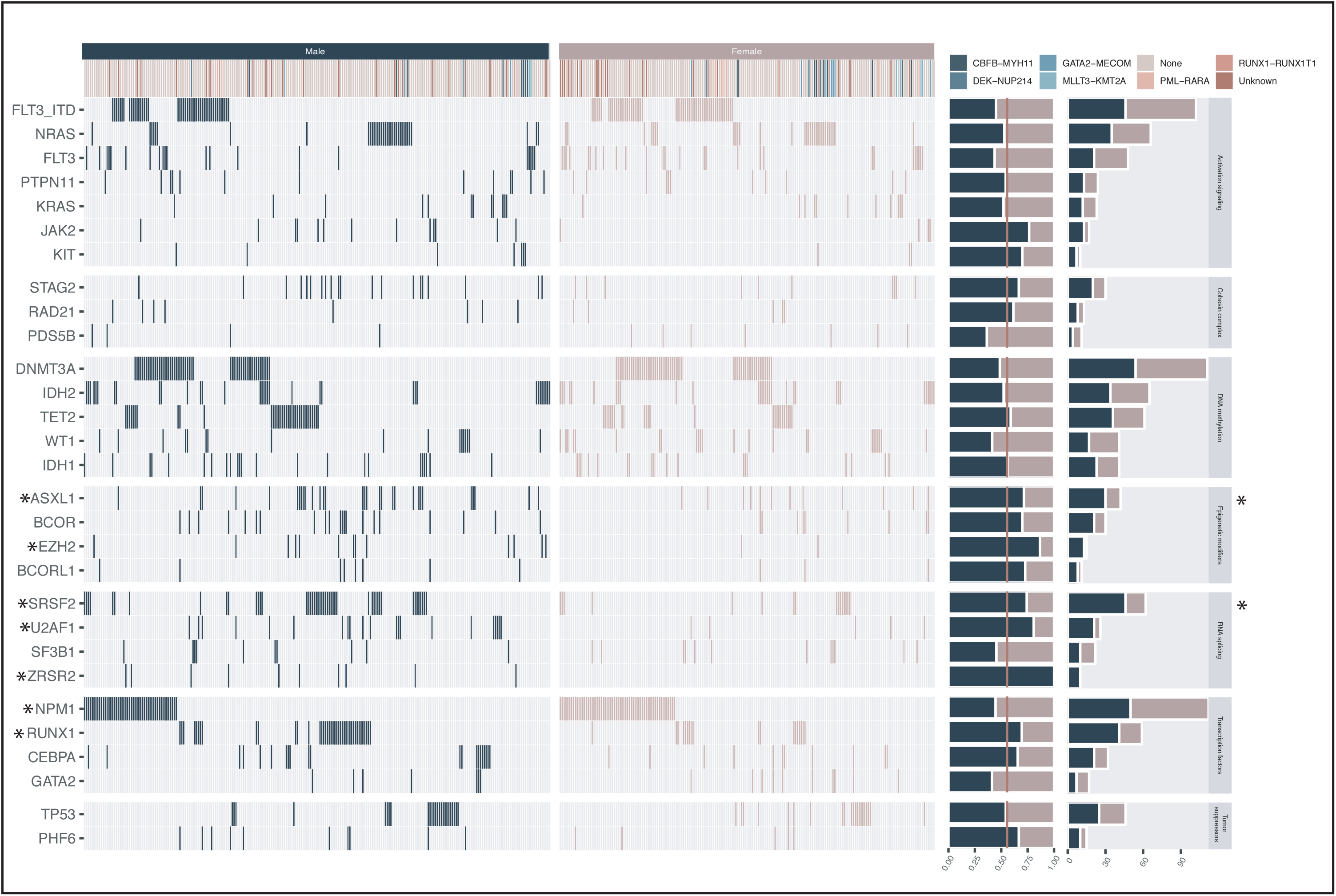
Sex-separated overview of genes mutated in more than 2% (≥10 patients) of the sub-sampled Beat-AML sample cohort (n = 498), identified by analysing the final curated exome sequencing variant list downloaded at http://www.vizome.org. Of note, the result of the *FLT3* gene is separated by ITD and non-ITD *FLT3* mutations on separate rows. In all other analyses in this report, *FLT3*-ITD annotations includes additional samples where *FLT3*-ITDs were identified exclusively by conventional methods.

The HOVON 1 and HOVON 2 cohorts comprise newly diagnosed AML patients aged 15-80 years treated on various study protocols of the Dutch-Belgian Hemato-Oncology Cooperative Group (HOVON) and the Leukemia Group of the Swiss Group for Clinical Cancer Research (SAKK) during the period 1987 to 2013. The sample selection analysed here was restricted to treatment-naïve non-APL AML patients with known *FLT3*-ITD status. This includes patients treated in the protocols HO04, HO04a,^17^ HO29,^18,19^ HO42,^20,21^ HO43^22^ and HO102.^23^ The studies are previously published, and sampling and data acquisition were performed as previously described.^24,25^ Patients included in HO102 are referred to as HOVON 2, while the remaining patients comprise HOVON 1. Detailed information on the individual trials is available at http://www.hovon.nl.

LAML-TCGA is a strongly selected sample cohort, composed to cover the major cytomorphologic and cytogenetic groups recognized in AML prior to 2013.^16^ The cohort comprises 200 de novo AML patients, including 92 females and 108 males ranging from 18-88 years. Due to its selective and unrepresentative composition, we have mainly used this cohort for integrated analyses. Single cohort comparative analyses of sex-differences related to clinical parameters were not performed. The TCGA-AML data analysis is based exclusively on variables as presented in the file “SuppTable01.xlsx”, downloaded from https://gdc.cancer.gov/node/876. For survival analysis, the variable “OS months 3.31.12” is used.

### Statistics

For comparison of continuous variables, we applied the non-parametric Wilcoxon rank sum test/Mann-Whitney test. The Fisher exact test was applied for comparison of categorical data. For DGE analysis, we log-transformed the CPM matrix provided in “Table S9-Gene Counts CPM” by the formula: CPM(log2+0.1). Analyses were performed in accordance with the Linear Models for Microarray Data pipeline in the Bioconductor R package (LIMMA version 3.38.3). For exploration of the drug sensitivity data from the Beat AML cohort, we analysed the area under the curve (AUC) values provided in “Table S10-Drug Responses”, applying categories from “Table S11-Drug Families”. Time-to-event analyses were performed by generation of Kaplan-Meier survival curves and compared for differences using the log rank test. For continuous variables, impact on outcome was evaluated by univariate cox models followed by multivariate logistic regression models. P-values were adjusted by the Benjamini-Hochberg method, and threshold was set at ≤0.05. All statistical analyses were performed in R-Studio (version 1.1.453) and R (version 3.5.0). Graphs were made with ggplot2 (version 3.1.0) and figures made in Adobe illustrator CS6 (version 16.0.0).

## RESULTS

### Comparative genomic architecture

To provide a context for sex-related variant patterns, we compared the sex-related distribution of somatic variants annotated in the Beat AML cohort. We found that the number of somatic variants did not differ significantly across sexes, although male individuals tended to have higher numbers (Supplementary figure 2A). Comparing the number of highly recurrently mutated genes (mutated in > 2% of the cohort), the same trend was apparent (Supplementary figure 2B). There were no sex-related differences within the *FLT3*-ITD mutated subgroup. We identified 28 highly recurrently mutant genes, of which 23 genes are autosomal and five are X-linked, including *BCOR, BCORL1, PHF6, STAG2*, and *ZRSR2*. The frequency of *FLT3* mutations and *DNMT3A* mutations were higher in females, although not statistically significant. However, seven other genes were significantly different; *NPM1* was overrepresented in females, while *RUNX1, ZRSR2, SRSF2, U2AF1, ASXL1* and *EZH2* were overrepresented in males (Figure 1). Notably, six of these seven genes are autosomal.

**Figure 2.**
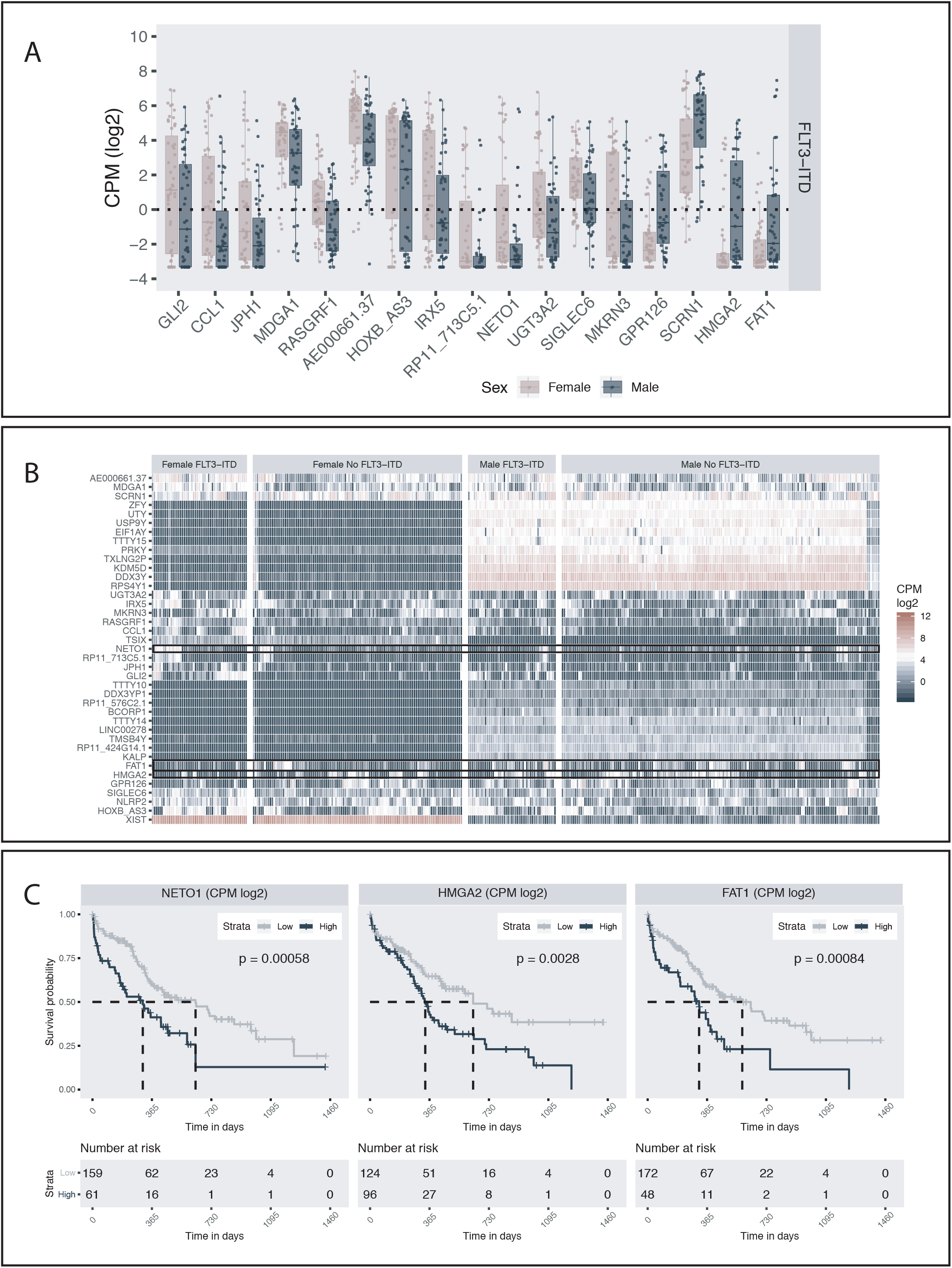
**A**. Pairwise comparison (Wilcoxon rank sum test/Mann-Whitney test) of the 17 transcripts identified as differentially expressed between female and male *FLT3*-ITD positive samples based on the results of the DGE analysis. Each dot represents an individual specimen and the box plot graphically present the median and the spread. The lower and upper hinges correspond the 25th and 75th percentiles, respectively, and the upper and lower whisker extends to the largest and smaller values (no further than 1.5 times the total inter quartile range from the hinges). Only *FLT3*-positive samples are presented. The same comparison for *FLT3*-ITD negative samples is available in supplementary figure 9. **B**. Heatmap depicting unsupervised clustering of CPM-log2 values of all transcripts significantly differentially expressed between female and male *FLT3*-ITD positive individuals. Clustering was done by columns and rows, and then manually faceted by *FLT3*-ITD status and sex. **C**. Kaplan-Meier curve comparing the outcome of patients characterised by high and low mRNA expression of *NETO1, HMGA2* and *FAT1*, respectively.

When expanding the selection to genes mutated in > 1% of the total Beat AML selection, we found mutations in *ZNF711* exclusively in females, while *BRCC3* and *SMC3* were both significantly overrepresented in males (Supplementary figure 4 and supplementary table 4A). Within the *FLT3*-ITD positive group, we identified mutations in both *BCOR* and *CCND3* only in male *FLT3*-ITD AML (Supplementary table 4B). Interestingly, the *FLT3*-ITD negative male and female groups were significantly different: while *WT1* was significantly more frequent in females, *RUNX1, SRSF2, U2AF1, ZRSR2*, and *EZH2* were significantly characteristic of males (Supplementary table 4C). This pattern is largely attributable to the excess of male samples annotated as “transformed” (from a prior hematological malignancy). Excluding these cases, only the relationship between male sex and *SRSF2* and *U2AF1* remained significant. In addition, *MXRA5* and *NF1* were identified as more frequently mutated in females, while *PHF6* was significantly overrepresented in males (Supplementary table 4D).

For comparison, the sex-related distribution of somatic variants in the LAML-TCGA cohort is presented in Supplementary table 5. Due to the selective composition of this cohort, integrated analyses were not performed.

We note that none of the significant sex-discrepancies observed in somatic mutations of single genes were statistically significant by adjusted p-value. However, many of our findings, including the association of *FLT3*-ITD, *NPM1* and *DNMT3A* mutations in females and *RUNX1, ASXL1, SRSF2, STAG2, U2AF1* and *EZH2* mutations in males, have previously been reported by others.^11–13^ Thus, we report all findings significant by non-adjusted p value here.

The 28 recurrently mutated genes in the Beat AML cohort were subsequently categorised by gene product functional properties: DNA methylation, activating signalling, transcriptional regulation, tumour suppressor function, epigenetic modification, cohesion complex regulation, and RNA splicing. We found that 14% of female vs 31% of male individuals had at least one mutation in one of the four RNA splicing genes: *SRSF2, U2AF1, SF3B1* and *ZRSR2* (adj. p = 0.0001). Similarly, 10% female vs 23% male individuals had at least one mutation in one of the four epigenetic modifier genes: *ASXL1, BCOR, EZH2* and *BCORL1* (adj. p = 0.0004). The relationships remained significant in the non-transformed group for both RNA splicing genes (adj p = 0.009) and for epigenetic modifier genes (adj. p = 0.03). The association was also significant within the *FLT3*-ITD negative group, where both epigenetic modifier genes (p = 0.01) and RNA splicing genes (p = 0.001) remained significantly associated with males. In the *FLT3*-ITD positive group, the same trend was apparent, although not significant by adjusted p-value (Supplementary table 6, Supplementary figure 5).

We further explored the distribution of mutations of the frequently *FLT3*-ITD co-mutated genes *DNMT3A* and *NPM1*. As this information was available for all four cohorts, integrated analysis was performed. Mutation status of all three genes was known in 1624 cases. We found that significantly more females than males had a mutation in at least one of these three genes. The combination *FLT3*-ITD and *NPM1* mutation without *DNMT3A* mutation, as well as the combination *FLT3*-ITD, *NPM1* and *DNMT3A* mutation were both significantly associated with females (Supplementary table 7, Supplementary figure 6).

We subsequently compared the distribution of variant allele frequencies (VAF) of the genes mutated at least 10 times in the Beat AML cohort, and found that VAF was significantly higher in male compared to female individuals in five genes. As expected, this included the X-linked genes *STAG2, PHF6* and *BCOR*, but also the autosomal gene *ASXL1. BCORL1*, despite being X-bound, did not have significantly higher VAF in males. *ZRSR2* was mutated exclusively in males (Supplementary figure 7).

*FLT3* was not among the genes identified with significantly different VAF across the sexes. It has presiously been shown that the allelic ratio (AR) of *FLT3*-ITD mutations has prognostic implications.^26,27^ To investigate whether there were sex-discrepancies in *FLT3*-ITD AR among these patients, we calculated the AR from the VAF (the approach is described in the figure legend of supplementary figure 8). We note that although there are no standardized approach to measure *FLT3*-ITD AR, it is commonly measured by DNA fragment analysis by capillary electrophoresis (CE).^28,29^ Here, we have used the VAF from next generation sequencing (NGS) anayses to calculate AR, as it has previously been shown that there is high concordance between CE and NGS assays in measuring *FLT3* mutational burden in AML patients.^30^ As was the case for VAF, we did not find significant differences in AR between males and females in this cohort (supplementary figure 8).

### Gene Expression Analysis

Based on the sex-related pattern of *FLT3*-ITD and co-mutations, we questioned whether there were sex-disparity in gene expression in the *FLT3*-ITD mutated group. We analysed 380 specimens in the Beat AML cohort, comprising 163 female and 217 male samples, respectively, of which 51 female and 47 male *FLT3*-ITD positive samples (supplementary table 8). We identified a total of 39 differentially expressed genes; 16 upregulated and 23 downregulated in the female *FLT3*-ITD positive subgroup, including 14 Y-linked, 2 X-linked, 7 non-annotated and 16 autosomal genes (Supplementary table 9). Subtracting genes also differentially expressed in the *FLT3*-ITD negative group, we identified a total of 17 mRNA transcripts mapped to 16 genes, of which *GLI2, CCL1, JPH1, MDGA1, RASGRF1, AE000661.3, HOXBAS3, IRX5, NETO1/RP11-713C5.1, UGT3A2, SIGLEC6*, and *MKRN3* were all significantly higher expressed in *FLT3*-ITD positive females, while *GPR126, SCRN1, HMGA2* and *FAT1* were significantly higher expressed in *FLT3*-ITD positive males (Figure 2A and B, Supplementary figure 9 and 10). Comparing *FLT3*-ITD positive (n = 98) and negative specimens (n = 282) irrespective of sex, we found that *GLI2, CCL1, JPH1, MDGA1, RASGRF1, AE000661.37, HOXB-AS3, IRX5*, and *RP11-713C5.1* were all significantly upregulated in *FLT3*-ITD positive AML, while *GPR126* was significantly downregulated.

To explore the functional relevance of genes identified as differentially expressed, we explored their inter-relationship with disease outcome in the 220 individuals in the Beat AML cohort where survival- and expression data was available. By univariate analysis, five of the 39 differentially expressed genes were identified as significantly correlated with outcome (Supplementary table 10): *NETO1, TMSB4Y, TTTY10, HMGA2* and *FAT1*. In multivariate analysis including these five transcripts, only *NETO1* remained significant, while *HMGA2* and *FAT1* were borderline significant (Supplementary figure 11). High expression of these three genes significantly correlated to poor outcome (Figure 2C). Stratifying the patients by *FLT3-*ITD status, we found that high expression of HMGA2 was significantly associated with poor prognosis in the *FLT3*-ITD negative sub-population only, whereas FAT1 and NETO1 remained significant in both *FLT3*-ITD positive and negative patients. Stratifying the patients by sex, high FAT1 and HMGA2 were significant negative prognostic markers in the male sub-population only, whereas high NETO1 was significant in both sexes, although a stronger negative correlation was observed in the male sub-population (Supplementary figure 12).

### Drug sensitivity and resistance testing

To further explore the functional consequences of the sex-discrepant leukemic architecture, we assessed the variation in drug sensitivity profiles in the Beat AML cohort (Supplementary table 11). We focused on the 113 compounds tested in a minimum of 100 specimens, and samples overlapping with our previous analyses, resulting in 348 specimens from 311 individuals. 96/113 compounds were annotated by mechanism of action and categorised into one or more of 39 different groups (Supplementary figure 13).

Comparing drug sensitivity across the 39 various groups between female and male individuals irrespective of ITD status, we identified a weak but significant relationship between RTK-RET inhibitors (1/39) and higher sensitivity in females. When comparing *FLT3*-ITD positive cases, six inhibitor families differed significantly: PI3K-AKT-MTOR, PI3K-MTOR, RTK-ERBB, RTK-INSR-IGF1R, SYK and CAMK inhibitors all demonstrated higher sensitivity in male samples (Supplementary figure 14). Comparing female and male *FLT3*-ITD negative specimens, only MEK inhibitors was significantly different, and more potent in female samples.

When examining individual compounds, we identified five compounds with significantly sex-discrepant potency, irrespective of *FLT3*-ITD status: Females were more sensitive to AT7519, palbociclib, and quizartinib (AC220), while males were more sensitive to cediranib (AZD2171), and pazopanib (GW786034). Further, we identified three compounds that were significantly more potent in female *FLT3*-ITD negative samples, including AT7519, palbociclib, and trametinib (GSK1120212). Importantly, we identified 14 inhibitors that demonstrated significantly lower potency in female *FLT3*-ITD positive samples, with an overrepresentation of inhibitors targeting tyrosine kinase receptors and downstream targets known to be influenced by FLT3-ITD signalling, including idelalisib, BEZ235, MGCD-265, masitinib (AB 1010), NVP-ADW742, tivozanib (AV-951), S31-201, afatinib (BIBW-2992), PRT062607, PI-103, cediranib (AZD2171), lapatinib, STO609 and axitinib (AG-013736) (Figure 3A, Supplementary table 12, Supplementary figure 15).

**Figure 3.**
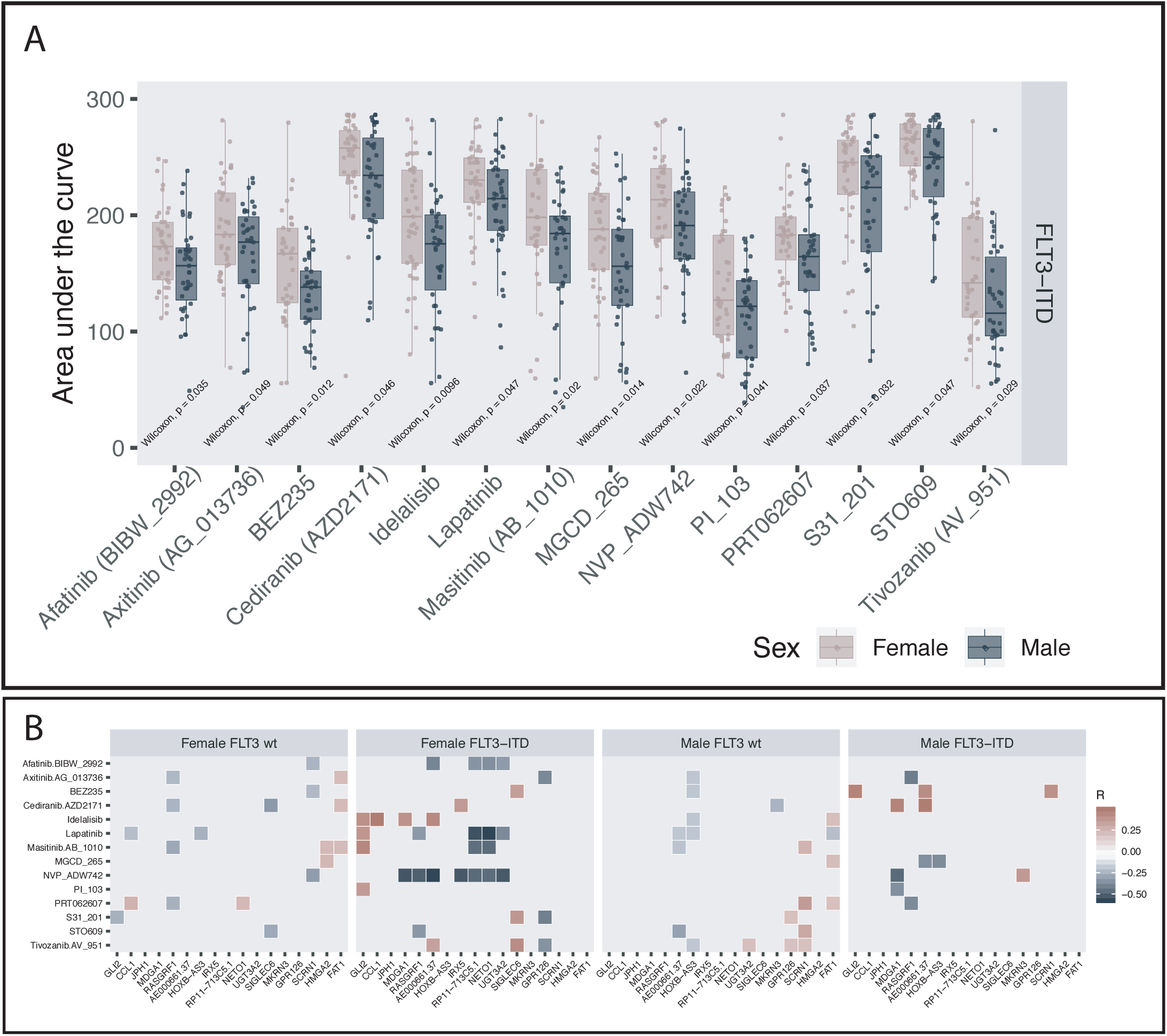
**A**. Comparison of area under the curve (AUC) between *FLT3*-ITD mutated female and male specimens for the 14 compounds identified to demonstrate significant sex-related divergence. The same comparison for *FLT3*-ITD negative samples is available in supplementary figure 13. (Wilcoxon rank sum test/Mann-Whitney test: p < 0.05). Additional statistical results for all compounds included in the analysis as well as and sample size for all compared groups are summarised in supplementary tables 12 and 13. **B**. Heatmap graphically presenting the pairwise correlation between the CPM-log2 values of the differentially expressed genes and the area under the curve distribution of compounds identified as differentially potent across female and male *FLT3*-ITD positive samples. The heat represents the correlation coefficient. Only statistically significant correlations are plotted.

To assess whether variation in gene-expression correlated with variation in drug-response, we further examined the pairwise relationships of the differentially expressed genes and the compounds with sex-discrepant potency in *FLT3*-ITD positive samples. We found that sensitivity to NVP-ADW742 correlated with increasing gene expression of 7/13 RNA transcripts identified as upregulated in female *FLT3*-ITD positive AML. Conversely, increasing *GLI2* expression correlated with reduced sensitivity to 4/14 drugs that were less potent in female *FLT3*-ITD positive AML (Figure 3B).

### Survival

Finally, we investigated the relationships between sex and the prognostic strength of *FLT3*-ITD mutation status on all cohorts combined as well as separately (Supplementary tables 14 a-e). HOVON 1 (n = 432) had a median follow up of 113.7 months while HOVON 2 (n = 625) had a median follow up of 42.3 months. The Beat AML cohort had a short median follow up of 15.2 months. For survival analyses, only newly diagnosed cases were included (n = 303). LAMLTCGA (n = 200) had a median follow up 47.2 months.

Despite *FLT3*-ITD status being a recognised negative prognostic marker in AML, we found a significant correlation to poor outcome only in the HOVON 1 cohort (Figure 4B). When overall survival was analysed in the cohorts combined (n = 1560), we found as expected that *FLT3*-ITD mutation status was associated with statistically lower survival (Figure 4A). However, when survival analysis for all cohorts combined was split by sex, the significant prognostic association of *FLT3*-ITD remained *only* in the female sub-population (Figure 4C). Analysing the cohorts independently, the same observation was apparent; *FLT3*-ITD was significantly correlated with poor outcome only in female patients in both the HOVON cohorts, and the same trend was observed in the Beat AML and LAML-TCGA cohorts, although not statistically significant (Figure 4D, Supplementary figure 16, Supplementary table 13 a-e).

**Figure 4.**
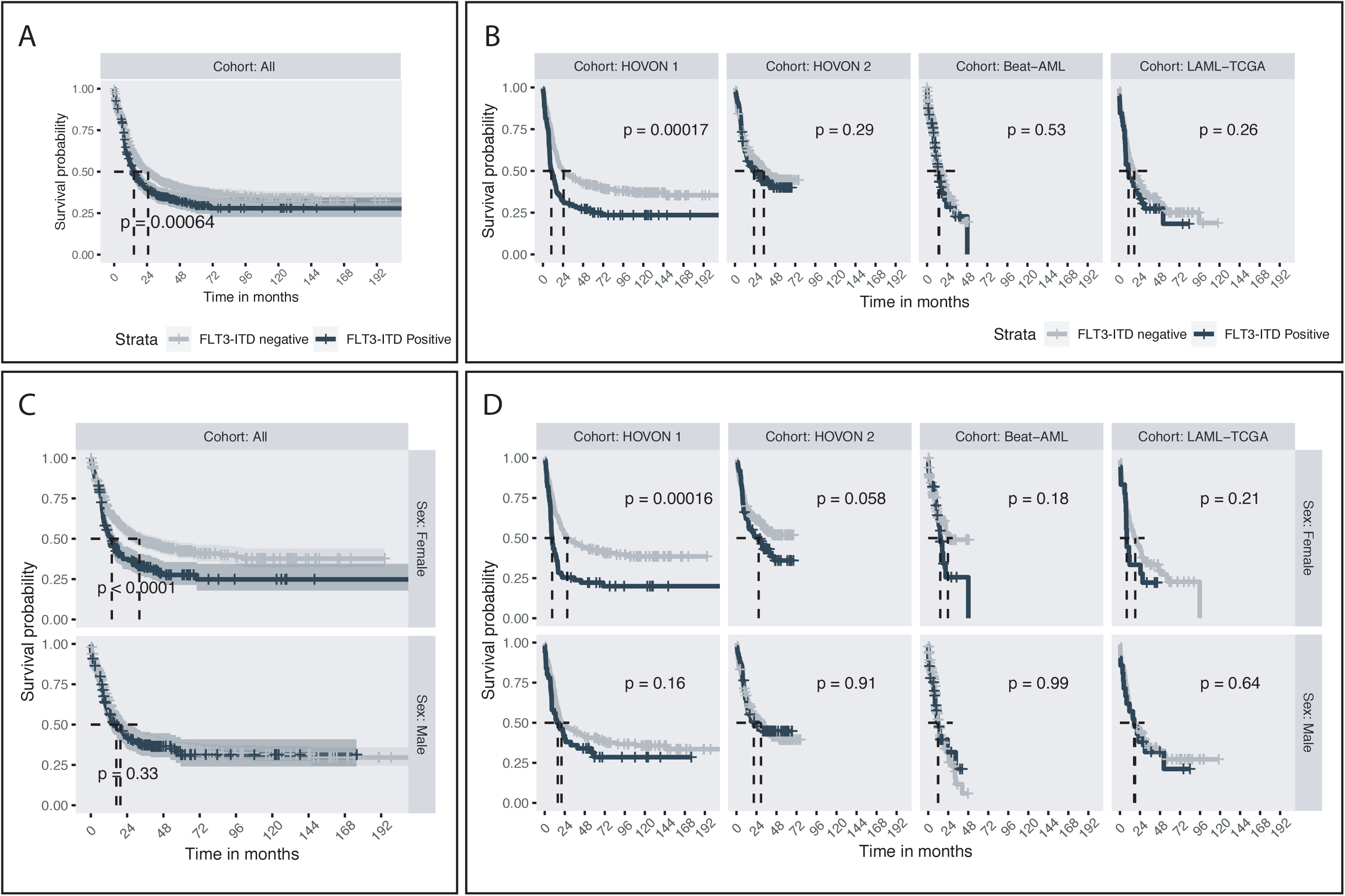
**A**. Kaplan-Meier curve comparing the outcome of *FLT3*-ITD mutated and non-*FLT3*-ITD mutated patients across the four cohorts analysed together. **B**. Kaplan-Meier curve comparing the outcome of *FLT3*-ITD mutated and non *FLT3*-ITD mutated patients analysed individually by cohort. **C**. Kaplan-Meier curve comparing the outcome of *FLT3*-ITD mutated and non *FLT3-*ITD mutated patients analysed across the four cohorts but analysed individually for females and males. **D**. Kaplan-Meier curve comparing the outcome of *FLT3*-ITD mutated and non *FLT3*-ITD mutated patients analysed individually by cohort and individually for females and males.

## DISCUSSION

In this work, we demonstrate sex-disparity in somatic variant composition, gene expression and *ex vivo* drug response patterns in *FLT3*-ITD mutated AML cases across four well-characterised AML cohorts. *FLT3*-ITD mutation status is integrated in standard risk stratification guidelines in AML. Yet, in the datasets we explored, *FLT3*-ITD mutation status separated the disease outcome only in female AML. This may indicate that the prognostic utility of *FLT3*-ITD mutation status is different across sexes. One plausible explanation is related to discrepant distribution of age, disease presentation pattern and poor risk molecular features, most pronounced in the Beat AML cohort. Here, the mutated genes dominating the male *FLT3*-ITD negative subpopulation are associated with poor outcome and with MDS and secondary AML.^31^ Whether this relationship represents inclusion asymmetry or a natural distribution is unknown. A characterisation of the population-based Swedish Acute Leukemia Registry, however, suggested that AML with an antecedent haematological disease is more frequent in males.^32^ Similar to AML, there is female excess of MDS among younger individuals, and men with MDS have comparably inferior outcome.^9^ Additionally, mutations in several genes overrepresented in males in the Beat AML cohort, including *SRSF2* and *U2AF1*, are reportedly overrepresented in male MDS.^33^

Another possible confounder is FLT3-ITD allelic ratio (AR). High AR has in several studies been linked to poor prognosis,^26,27^ and it has also been shown that AML patients with a high *FLT3*-ITD AR lacking *NPM1* mutations have a worse prognosis.^34^ In the latter study, an excess of males was reported in this sub-group, although no significant sex-disparity was observed. In our analyses, we did not identify *FLT3*-ITD VAF nor AR as significantly different across sexes, suggesting that a difference in the mutational burden of *FLT3* is not the cause of the observed sex-discrepancy in the prognostic impact if *FLT3*-ITD.

The sex-specific age-distribution characterising AML demography suggest that cohort composition is an important confounder. The age-composition of the Beat AML cohort resembles the reported demographic distribution of AML far better than the strongly selected LAML-TCGA cohort, in part explaining the lack of sex-biased mutations in this cohort. One could argue that age-matching is the optimal condition for comparison. What remains, however, is to identify the cell-intrinsic and cell-extrinsic mechanisms underlying the sex-disparity in AML incidence and molecular presentation. Mutations are stochastic events; thus there are no apparent reasons to suggest that male and female hematopoietic stem cells acquire discrete mutations. What is plausible, however, is that sex is an independent contextual contributor in determining the translational effect in comparative fitness of novel gene variants.

Interestingly, one of the very first papers characterising the FLT3 receptor suggested FLT3 mediated regulatory function not only in the hematopoietic compartment but also in the gonads, placenta and brain;^35^ tissues characterised by sexual dimorphism. This suggests that downstream effects of FLT3 signalling may be influenced by sex-variation. This hypothesis has been substantiated by studies demonstrating functional relevance of FLT3 expression in germinal tissue^36,37^ and by investigation of the oestrogen receptor in FLT3 positive haematopoietic cells, progenitor cells and mature cellular subsets like dendritic cells.^8,38^ Together, these observations suggest inter-regulatory pathways between sex-steroid receptors and FLT3.

The DGE analysis showed distinct expression of leukemogenesis-associated genes, suggesting functionally relevant cell-intrinsic sex-dependent differences. Of the genes more highly expressed in female *FLT3*-ITD positive AML, the hedgehog signalling mediator *GLI2* has been associated with *FLT3*-ITD positive leukemia,^39^ while *HOXB-AS3* is reportedly upregulated in *NPM1* mutated AML, and has been implied to contribute in regulation of AML cell-cycle progression.^40^ Of the genes more highly expressed in male *FLT3*-ITD positive AML, *FAT1* was shown to be somatically mutated in *FLT3*-ITD positive AML, and significantly in combination with *NPM1* and *DNMT3. FAT1* was further shown to exert tumour-suppressor activity specifically in *FLT3*-ITD positive AML,^41^ perhaps providing a partial explanation for the lack of prognostic impact of *FLT3*-ITD in this subpopulation. The negative prognostic impact related to differentially expressed genes was stronger in genes specifically down-regulated in female *FLT3*-ITD positive AMLs, namely *HMGA2* and *FAT1*. This may suggest that the sex-related discrepancies of outcome in *FLT3*-ITD mutated AML could be influenced by discrete molecular mechanisms.

We hypothesize a significant contribution of sex-specific leukemia-host interactions related to *FLT3*-ITD mutations. *Ex vivo* drug screens have shown that *FLT3*-ITD dependency is contingent on availability of growth factors, and response to FLT3-targeting drugs is partly permissive of culture conditions.^42^ This is substantiated by observed sex-related *ex vivo* drug response patterns in the study of the Cancer Cell Line Encyclopedia.^43^ An important confounder for *ex vivo* drug screens is incubation time, and conclusions regarding the translational value of these data should be drawn with caution. Nevertheless, the sum of the *ex vivo* drug response patterns and clinical outcome analysis suggest that response to therapeutic intervention in *FLT3*-ITD mutated AML may be influenced by sex. This is in line with the observations in the phase III trials RATIFY^14^ and QuANTUM-R^44^ treating *FLT3* mutated AML with midostaurine and quizartinib, respectively, both reporting survival benefit in the male sub-population only. Sex-divergent responses have also been reported in tyrosine kinase inhibitor treatment with sunitinib.^45^ These observations may be of great importance for the field of precision hematooncology, as novel targeted therapies may benefit female and male individuals differently. Although men and women in an unstratified AML population have similar prospects, this picture may be significantly different within discrete molecularly defined strata.

## CONCLUSIONS

Assessment of mutation status is becoming an integrated part of the diagnostic characterization of adult AML patients. This information is subsequently incorporated into risk stratification models and clinical decision-making. *FLT3*-ITD mutation status is well-established as a biomarker in AML. However, important questions remain regarding its optimal application and utility. Our observations suggest that *FLT3*-ITD mutation status should be optimized as a clinical tool in a sex-adjusted manner. Furthermore, we suggest that sex-specific considerations should be included in clinical risk stratification and considered in pre-clinical and clinical experimental design and in biomarker analyses in AML. Sex should further represent an independent stratification factor when randomizing to clinical trials, and be systematically included when analyzing and reporting on clinical data in AML. We hypothesize that addressing sex-related regulation of molecularly defined subgroups of AML could advance pathophysiological understanding, perhaps ultimately revealing new therapeutic possibilities.

## Data Availability

The data from the BeatAML and TCGA sample cohorts analysed are exclusively from publically available datasets. All results from statistical analyses are included in the supplementary material.

http://www.vizome.org/aml/geneset/

https://gdc.cancer.gov/node/876

## ACKNOWLEDGMENTS

We are grateful to all patients that have participated in the respective clinical trials, and all members and clinical investigators of the HOVON/SAKK collaboration, the Cancer Genome Atlas and in the preparation and publication of the Beat AML sample cohort.

## AUTHORSHIP

The study was designed by CE and BTG. CE analysed, interpreted, prepared data for presentation and wrote the paper. MH and BTG interpreted data and wrote the paper. TG, BL and PJMV collected data from the HOVON cohorts, and contributed with data interpretation and with manuscript preparation.

